# Immunosuppression as a hub for SARS-CoV-2 mutational drift

**DOI:** 10.1101/2022.06.09.22276203

**Authors:** Guy Shapira, Tal Patalon, Sivan Gazit, Noam Shomron

**Affiliations:** Faculty of Medicine, Tel Aviv University, Tel Aviv, Israel; Edmond J. Safra Center for Bioinformatics, Tel Aviv University, Tel Aviv, Israel; Kahn Sagol Maccabi (KSM) Research & Innovation Center, Maccabi Healthcare Services, Tel Aviv, Israel; Maccabitech Institute for Research and Innovation, Maccabi Healthcare Services, Israel

## Abstract

Newly emerging SARS-CoV-2 variants of concern (VOCs) play a major role in the persistence of the coronavirus disease 2019 (COVID-19) pandemic. While these VOCs are characterized by extraordinary evolutionary leaps and evasion from previously acquired immunity, their origins remain mostly unknown. In this study, we paired electronic health records of individuals infected with SARS-CoV-2 to viral whole-genome sequences, to assess effects of host clinical parameters and immunity on the intra-host evolution of SARS-CoV-2. We found small, albeit significant differences in SARS-CoV-2 intra-host diversity, which depended on host parameters such as vaccination status and smoking. Viral genomes showed a significant difference in the early course of disease in only one of 31 immunosuppressed patients, a recently vaccinated woman aged in her 70s. We highlight the unusually mutated viral genome obtained from this woman, which harbored near-complete truncating of the accessory protein ORF3a. Our findings suggest only minor influence of host parameters on the SARS-CoV-2 intra-host evolutionary rate and trajectory, with even the majority of immunosuppressed persons carrying fairly unremarkable viral genomes. We hypothesize that major evolutionary steps, such as those observed in VOCs, are rare occurrences, even among immunodeficient hosts.

**Highlights:**

- Intra-host viral diversity is modestly affected by host clinical parameters, such as vaccination status and smoking.
- The emergence and fixation of high-impact SARS-CoV-2 mutations in immunosuppressed hosts are rare, and not exclusive to patients with prolonged viral shedding.
- We identified a rare stop-gain mutation, leading to near-complete truncating of ORF3a, in an immunosuppressed woman recently vaccinated against COVID-19.

## Introduction

The continued emergence of new variants of concern (VOCs) of severe acute respiratory syndrome coronavirus-2 (SARS-CoV-2) is a persistent global health threat. Over the course of the pandemic, we have observed VOCs with increased pathogenicity, infectivity and immune escape capabilities, each more likely to overcome previously-acquired COVID-19 immunity^1^. These VOCs are inconsistent with the relatively low evolutionary rate observed in circulating lineages (roughly 5-fold lower than in influenza), and thus represent evolutionary leaps that cannot be explained by the gradual accumulation of mutations observed in standard conditions^2^. Current theories for the elusive origin of new variants suggest either prolonged infection in an immunodeficient human host or spillover infections in susceptible animals, or a combination of the two^3^.

Intra-host SARS-CoV-2 mutations emerge and accumulate during the course of infection. While mostly stochastic, recent studies have revealed distinct mutational patterns that are significantly affected by clinical parameters of the host^4^. Upon infection, SARS-CoV-2 is subjected to positive selection, giving rise to many, largely nonsynonymous mutations. Upon transmission, the virus is subjected to purifying selection, and most nonsynonymous intra-host single nucleotide variants (iSNVs) are not transmitted to new hosts^5^.

Immunosuppressed SARS-CoV-2 hosts are at risk of persistent viral shedding, which lasts several months and accumulates mutations that contribute to immune evasion^6^. To investigate the effects of immunosuppression and other clinical parameters on the intra-host evolution of SARS-CoV-2, we examined whole genome sequences of SARS-CoV-2 in relation to the electronic health records of their hosts.

## Results

### Data overview

The cohort is composed of 1,304 Israeli COVID-19 patients, diagnosed between March 27, 2020 and July 22, 2021, who had a positive SARS-CoV-2 polymerase chain reaction (PCR) test, as well as whole genome sequencing data of the virus that infected them (Table 1). A single SARS-CoV-2 sample was collected from each of the included persons.

**Table 1:** Characteristics of the cohort. ^a^Clinical and lifestyle parameters at the diagnosis of SARS-CoV-2 infection. S.E.: standard error; N.A.: not available

| Group | Immunocompetent<br>(N=1,273) | Immunosuppressed<sup>a</sup><br>(N=31) |
| --- | --- | --- |
| Males (fraction) | 692/1,273 | 20/31 |
| Age, mean ± SE | 35.6 ± 0.5 | 60.8 ± 3.1 |
| Diabetes mellitus<sup>a</sup>, % | 7.20% | 41.90% |
| Vaccinated<sup>a</sup>, % | 90.40% | 96.70% |
| Smoker<sup>a</sup>, % (N.A. %) | 10.4% (24%) | 6.4% (0%) |

### Distribution and characteristics of SARS-CoV-2 mutations

To classify viral mutations as either intra-host (emerging in a host over the course of infection) or inter-host (transmitted from a previous host), we relied on the type of mutation and its relation to phylogenetically similar viral sequences. In short, an iSNV was defined as a SNV with a low allele frequency (present in less than 90% of the reads) that did not appear in neighboring viral sequences. SNV with high allele frequency that were sufficiently rare and absent from other viral sequences in the clade, was also considered iSNV (see the Methods).

The abundance of iSNVs differed between viral clades; the delta variant showed lower intra-host diversity than did the alpha variant (mean±SE 3.6±0.4 vs. 11.1±0.9 iSNVs per sample). For both viral clades, iSNVs decreased in diversity as the viral population reached stability (fig 5). These global changes in the SARS-CoV-2 mutational profile led to an overall decrease in intra-host diversity, which was associated with time (P<2e-8; Z=-5.6) and with the gradual replacement of the alpha variant with the delta variant (P<7e-13; Z=-7.1).

Considering the aforementioned factors, iSNVs are significantly more abundant in vaccinated hosts (P<1e-7;Z=5.3) and older persons (Z=4; P<6e-5), and significantly less abundant in current smokers (Z=-4; P<4e-6). Immunosuppressed hosts and persons with diabetes showed a slight, albeit insignificant decrease in overall iSNVs (Z=-1∼; P=0.1 and P=0.16, respectively). Associations were not found of iSNVs with a patient’s sex or disease severity (P>0.2).

### The identification of an unusual viral genome from an immunosuppressed host

While no significant differences were found in intra-host variability between samples acquired from immunosuppressed and immunocompetent hosts, one sample had a highly unusual viral genome, harboring many iSNVs. We highlight a SARS-CoV-2 genome obtained from an immunosuppressed woman aged in her 70s who tested positive for infection 37 days after receiving a second dose of the BNT162b2 vaccine, during a Delta-dominant period in Israel. She has been immunosuppressed since 2014, and has minimal change disease and Hashimoto’s thyroiditis. Despite her medical conditions, she recovered from COVID-19 within one month without the need for hospitalization, and had no record of COVID-19-related symptoms, except for post-infection fatigue.

Analysis of the woman’s viral genome sequence revealed a large excess of rare, nonsynonymous mutations. In particular, ORF1ab contained 10 missense mutations that were unique within our dataset and also extremely rare globally.

We identified a stop gain mutation in the beginning of the ORF3a gene, leaving only the first 10 amino acids of a total 275 in the ORF3a protein (G25423T; ORF3a:p.11G>*) (Table 2). The mutation is completely fixed (present in all 3,117 reads covering the locus) and was not identified elsewhere in the dataset; the mutation was reported in only two other samples (according to GISAID; 10/4/2022). Due to the fixation of the mutation and the location of the premature stop codon upstream of all functional domains, complete loss of function in ORF3a is expected.

**Table 2:**
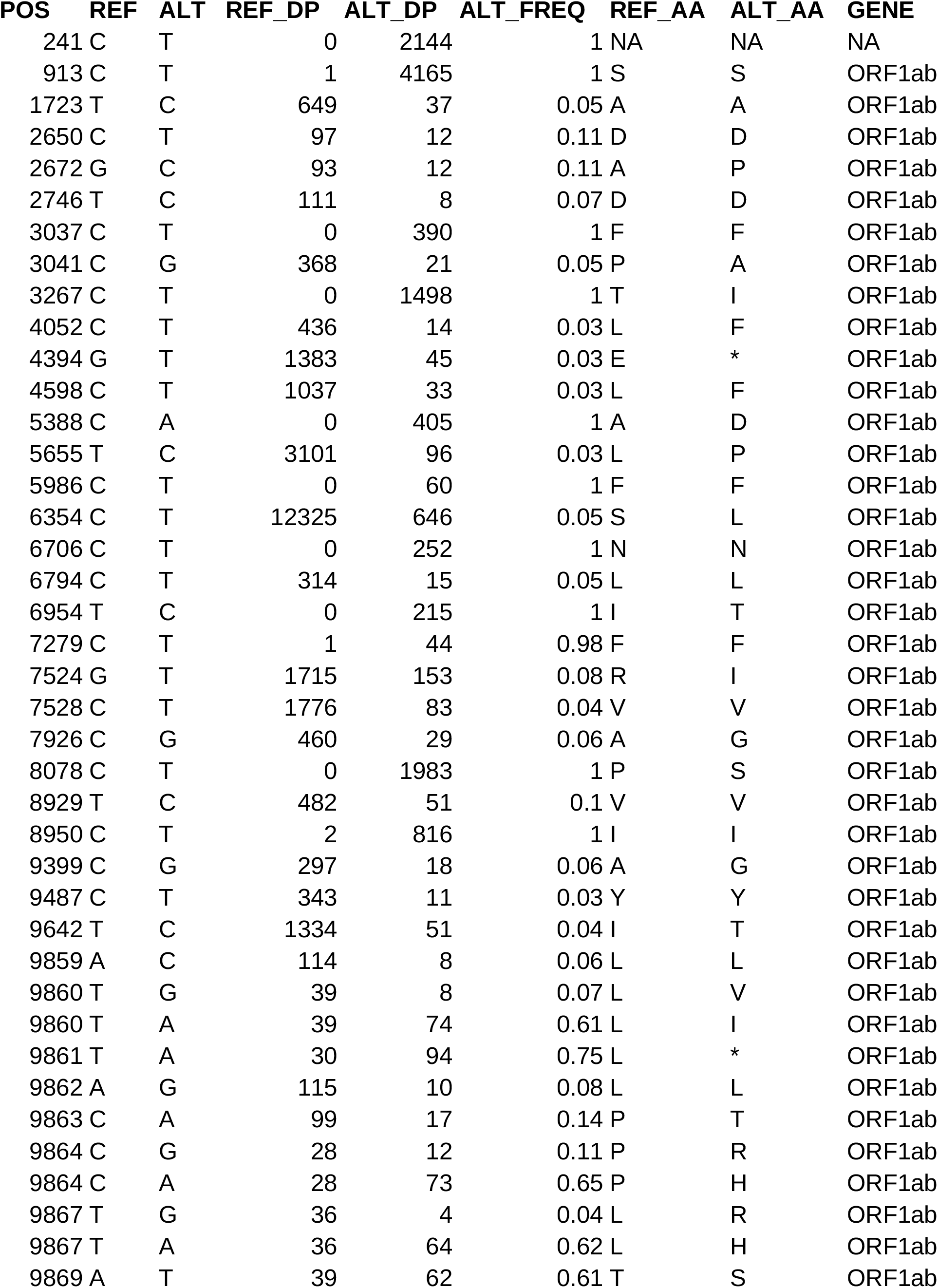
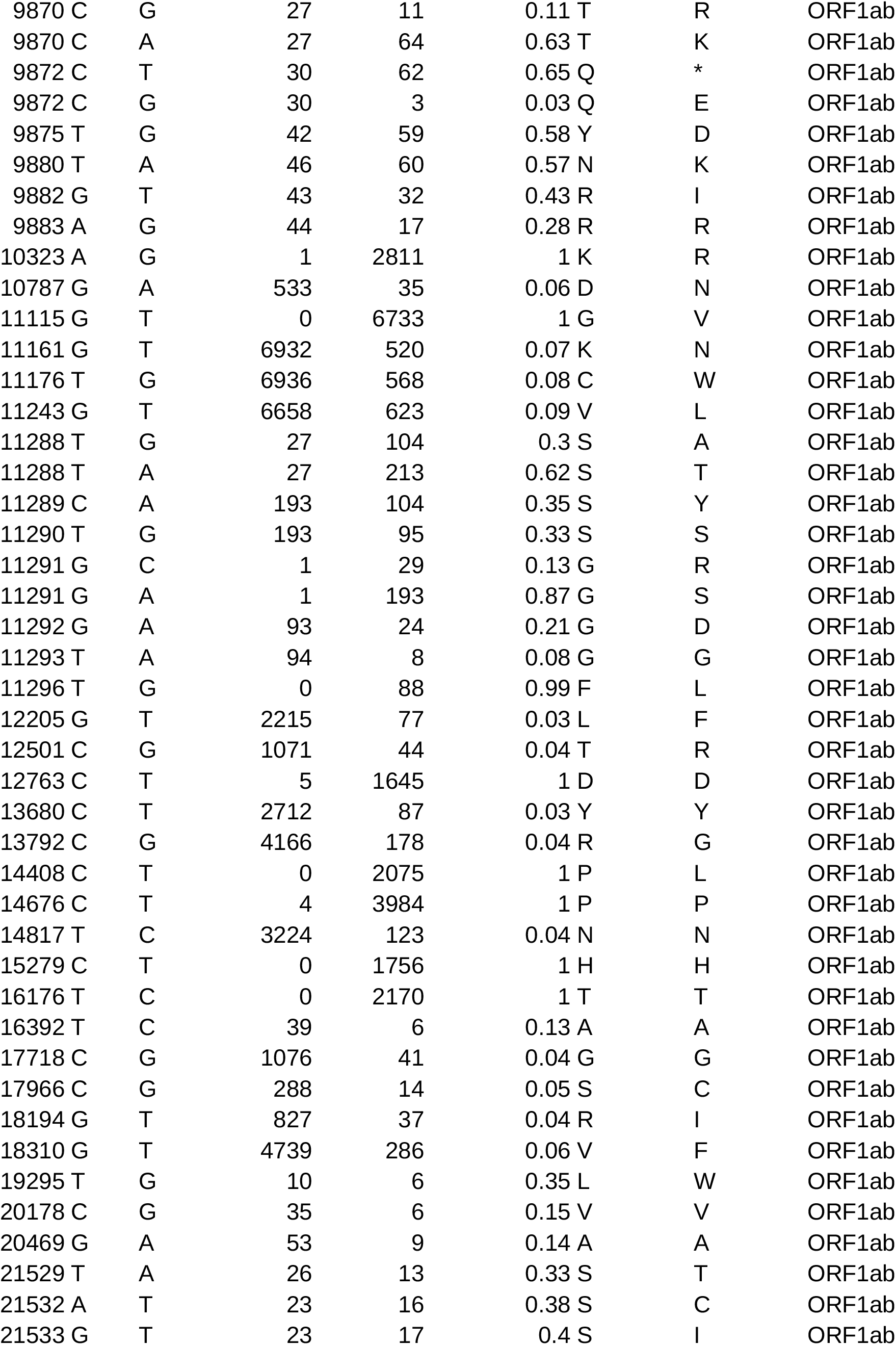
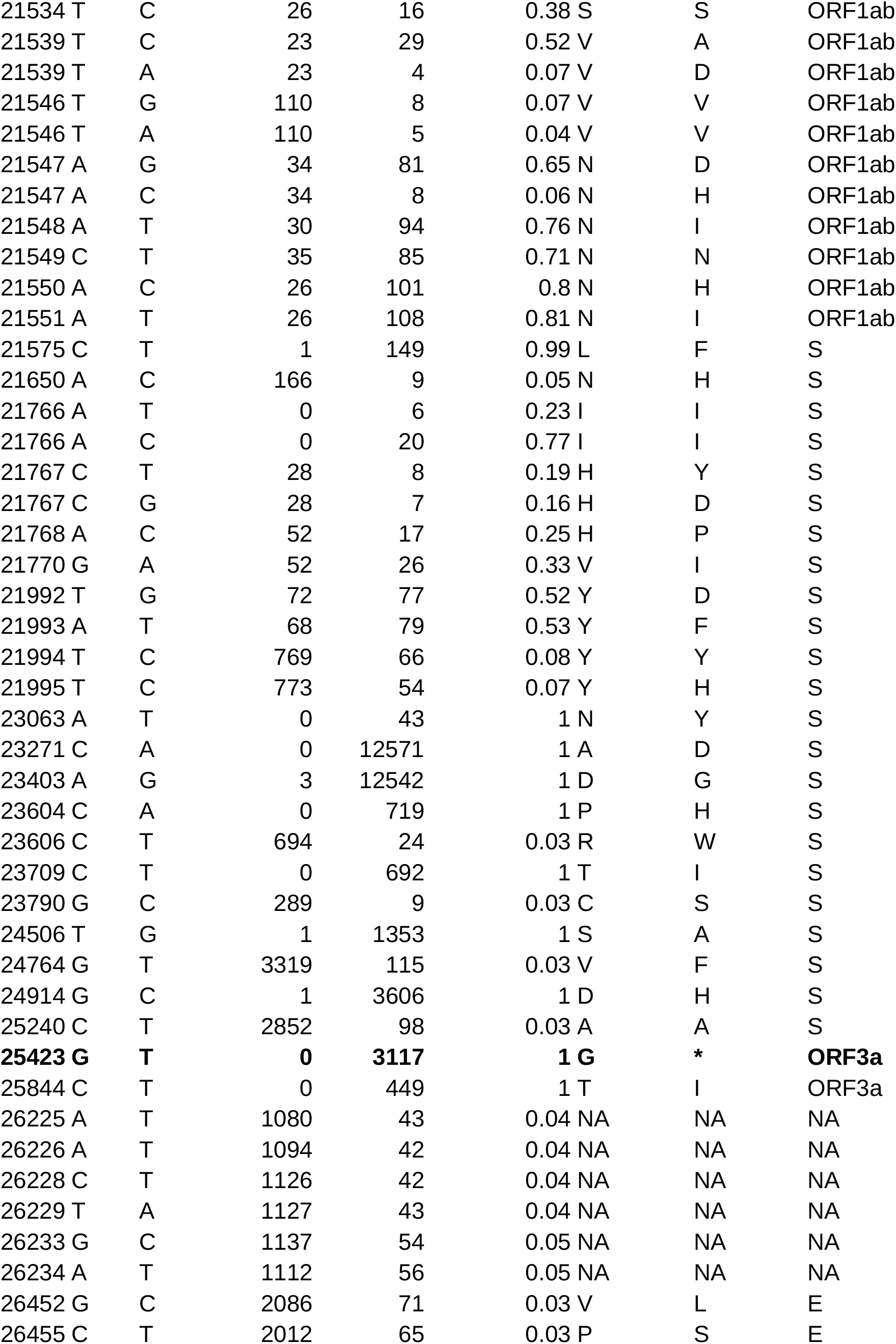
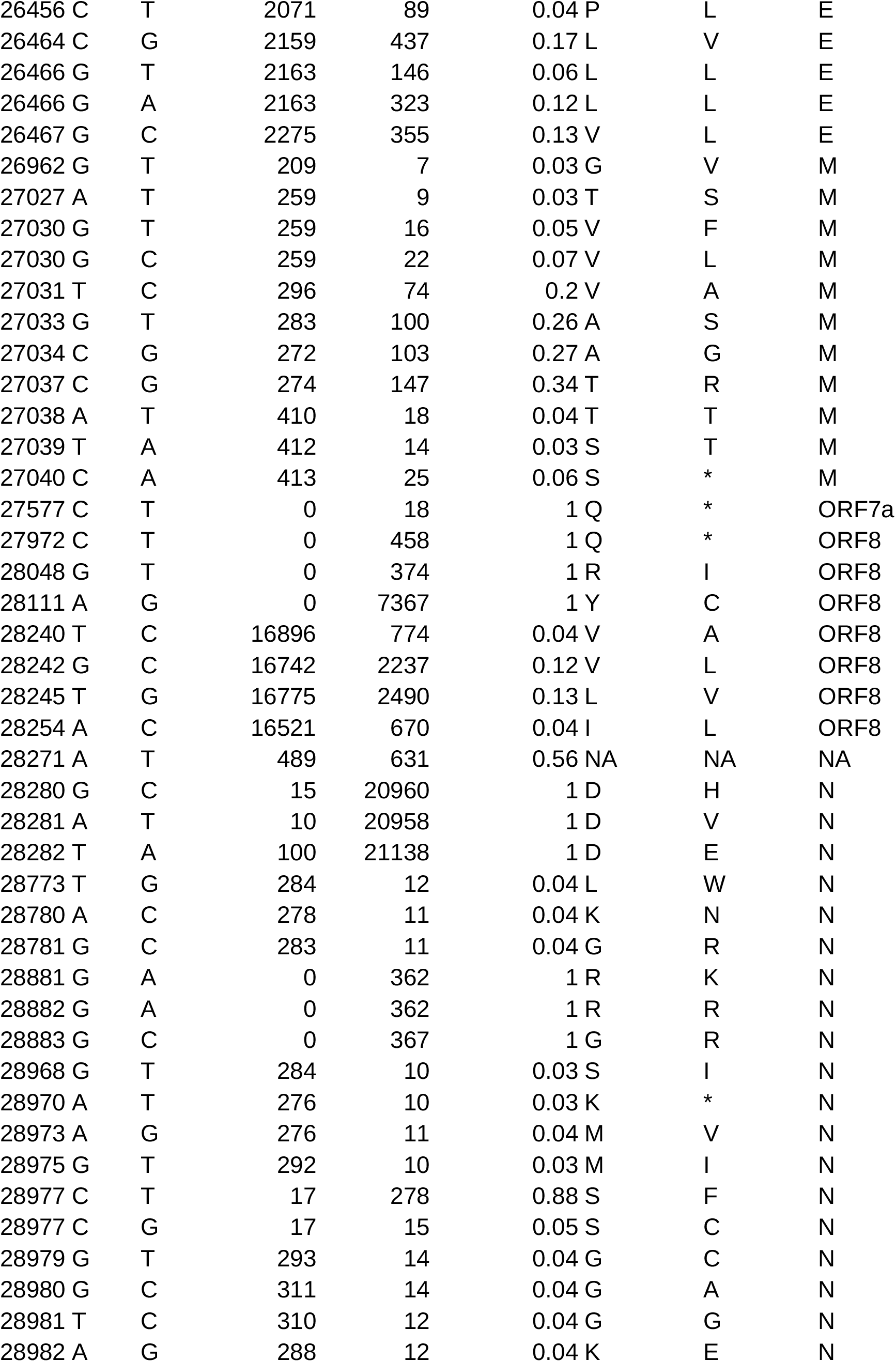
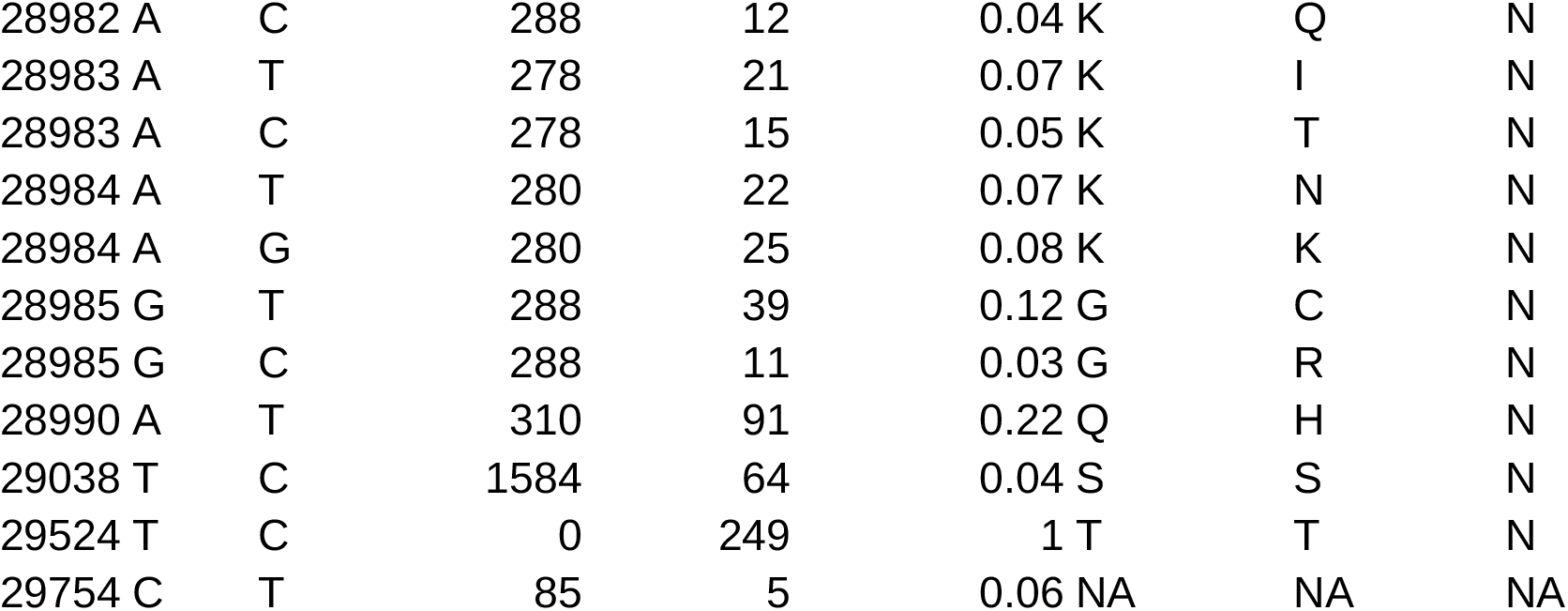
All the mutations found in the SARS-CoV-2 genome sampled from the immunosuppressed woman who harbored the ORF3a stop gain mutation (the ORF3a mutation is highlighted).

## Discussion

### A weak association between SARS-CoV-2 intra-host variation and host factors

Our analysis of SARS-CoV-2 intra-host diversity suggests slight differences in the abundance of iSNVs, relative to a host’s age, vaccination status and smoking habits. The relatively weak association between host factors and the intra-host mutation rate is consistent with previous findings. However, our analysis indicates a positive correlation with age, while the longitudinal study by Li and colleagues found a slight negative correlation^4^.

We estimate that the intra-host differences observed in mutational rate might be indicative of differences in host-pathogen interactions, yet the transmissibility and impact of these differences on overall viral evolution is likely limited, owing to strong purifying selection^7^. The nonsynonymous iSNVs were almost exclusive to the lower end of allelic frequency (Figure 1) and no evidence was found of fixation of any iSNVs in the cohort in the wider viral population.

**Figure 1:**
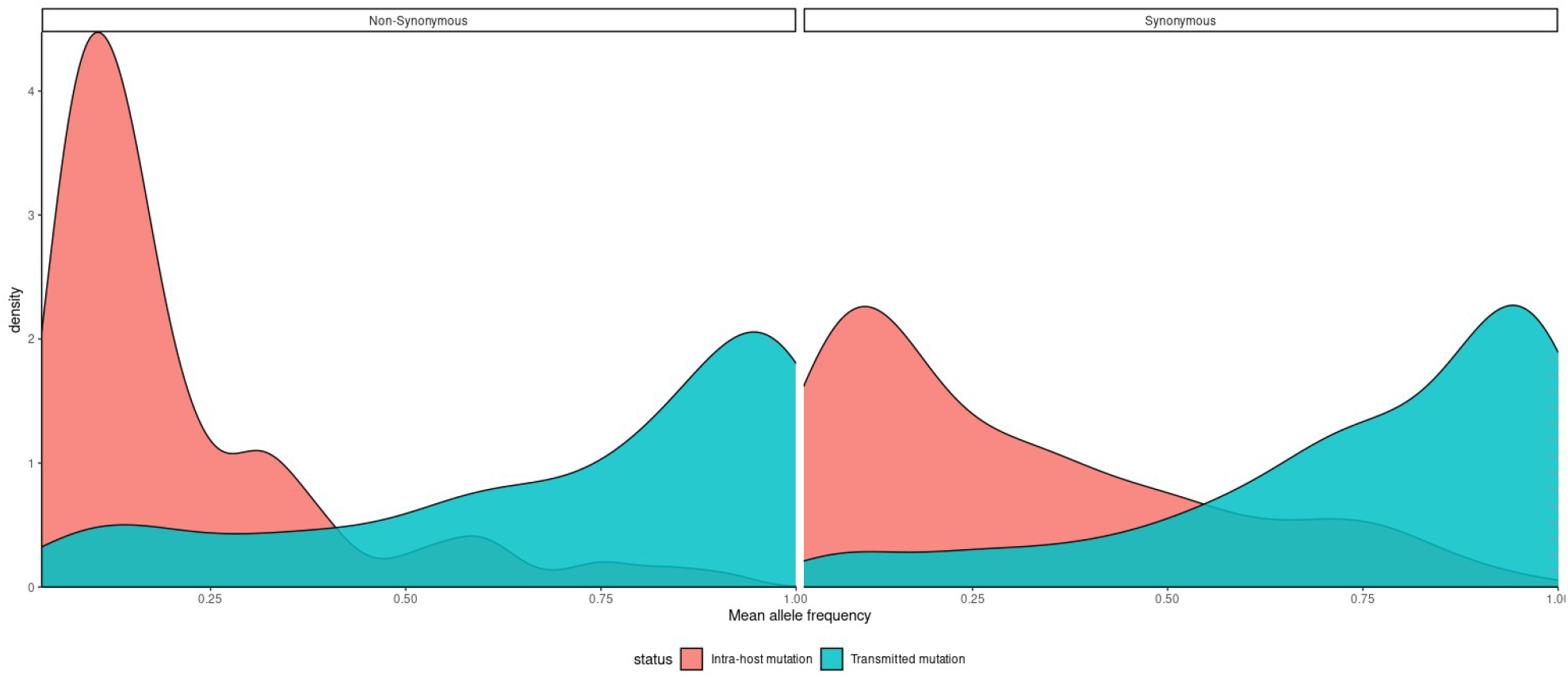
A density plot of transmitted and non-transmitted (intra-host) mutations by their mean allele frequencies, stratified to synonymous (left) and nonsynonymous (right) mutations.

### Immunosuppression and SARS-CoV-2 evolution

Surprisingly, viral genomes from immunosuppressed hosts did not differ significantly from those derived from immunocompetent hosts. Notably, our immunosuppressed cohort was almost entirely fully-vaccinated; the samples were taken early over the course of infection and none of them showed prolonged viral shedding. It is well established that immunosuppressed vaccine recipients have significantly lower seroconversion rates^8^, but the highly variable humoral response observed suggests that this group cannot be generalized without prior knowledge of antibody titres^9^. This raises the possibility that our immunosuppressed cohort had a largely standard course of SARS-CoV-2 infection, both in duration and intra-host evolution.

A single immunosuppressed woman, who had Hashimoto’s thyroiditis and minimal change disease, harbored a highly unusual viral genome. She was infected roughly one month after the second vaccine dose, when antibody levels typically reached their peak among the general population^10^. An excessive amount of rare missense mutations was found to affect ORF1ab and complete truncation of ORF3a past the tenth codon, which we named ORF3a^Δ^. A gradual accumulation of unusual SARS-CoV-2 mutations was previously reported in immunosuppressed hosts after months of prolonged viral shedding^11^, but in the woman described here, the mutations occurred early in the course of infection.

### ORF3a truncation in an immunosuppressed host

Truncating mutations were previously reported in SARS-CoV-2 accessory proteins, such as ORF7a and ORF8^12,13^. However, the truncated portion of ORF3a that we identified is much larger and located directly upstream to one of the most conserved loci in the viral genome, where the occurrence of a loss of function mutation is not as common^14^. As the premature stop codon is upstream from all functional domains, with the remaining peptide cleaved from the protein product, ORF3a^Δ^ is predicted to cause complete loss of function of the protein. Two out-of-frame ORFs were identified downstream of ORF3a^Δ^; however, these cannot compensate for the truncation of the canonical ORF, as the peptides produced are non-functional^15^. Due to the rarity and predicted deleteriousness of the mutation, combined with the overall unusual characteristics of this viral genome, we hypothesize that the mutation occurred within the host and that its emergence can be attributed to the immunosuppressed state.

ORF3a is the largest SARS-CoV-2 accessory protein, and has a diverse array of functions in the host cell, including subcellular localization to the plasma membrane and golgi, induction of apoptosis and inhibition of interferon signaling^16–19^. ORF3a induces incomplete autophagy, then subverts the formation of the autolysosome^20,21^. In addition to the functions of the canonical ORF3a protein, three overlapping protein-coding genes (named ORF3b, ORF3c and ORF3d) were more recently identified and found to contribute to immune evasion; however, the extent of their functions remains mostly unknown^22^.

Nemudryi et al. identified a truncating mutation in ORF7a that eliminated translation of a viral transmembranal domain, and that limited replication and viral immune suppressoin^12^. Like ORF3a^Δ^, the ORF7a truncation is hypothesized to have originated from an immunosuppressed host and to have limited spread. Unlike ORF3a^Δ^, the ORF7a truncation circulated in a small local community for about 1.5 months before disappearing. Based on this comparison and the importance of ORF3a to the viral process, we hypothesize that ORF3a^Δ^ is exceptionally deleterious, possibly to the point of being endemic to severely immunodeficient hosts.

Unlike the slow accumulation of immunity-evasive spike mutations that have been observed in immunosuppressed hosts^11^, ORF3a^Δ^ is a rapidly fixated, maladaptive mutation that is likely to impair viral fitness. This largely unknown class of mutations is rarely observed and the extent of its influence on SARS-CoV-2 evolution is unclear.

## Conclusion

The preliminary evidence presented here suggests mild associations between viral iSNVs and host factors, such as vaccination and smoking. In addition, we highlighted an unusually mutated viral genome, which includes a complete loss of function mutation in ORF3a that emerged in an immunosuppressed host shortly after initial SARS-CoV-2 infection. Our findings suggest that profound SARS-CoV-2 evolutionary changes are rare even among immunosuppressed hosts, but might occur relatively early over the course of infection. The association between intra-host evolution and basic clinical parameters remains uncertain and requires further research.

## Materials and methods

### Data sources and collection

#### Data sources

Anonymized electronic health records were retrieved from the centralized computerized database of Maccabi Healthcare Services (MHS). MHS is a state-mandated, non-for-profit, health fund in Israel that insures and provides healthcare services to 26.7% of the Israeli population. The MHS computerized database has been maintained centrally for over thirty years. This enabled a comprehensive longitudinal medical follow-up, including demographic data, clinical measurements, outpatient and hospital diagnoses and procedures, medications dispensed, imaging performed and comprehensive laboratory data from a single central laboratory.

#### Data collection

Demographic data included sex, age and a coded residential socioeconomic geographical statistical area, assigned by Israel’s Central Bureau of Statistics, which is the smallest geostatistical unit of the Israeli census (corresponding roughly to neighborhoods) and which embodies socioeconomic status. COVID-19 related information consisted of vaccination dates and the results of any PCR tests for SARS-CoV-2. COVID-19-related hospitalizations and mortality records were retrieved as well. COVID-19 related data also included viral sequencing of some of the PCR tests, which were chosen randomly by MHS. Additionally, data comprised information on chronic conditions from MHS’s automated registries, including immunocompromised conditions, cardiovascular diseases, hypertension, diabetes mellitus, chronic kidney disease, chronic obstructive pulmonary disease and obesity (defined as a body mass index of 30 kg/m2 or higher). The study population included all MHS members who had both a positive PCR test result and viral sequencing information of that test.

### Processing of whole-genome viral sequencing data

Raw sequencing data were trimmed using fastp 0.21.0^23^, aligned with bwa-mem2 2.2.1 to the MN908947.3 SARS-CoV-2 reference assembly, and then underwent primer trimming (all the samples were prepared with Artic V3 primers), variant calling and consensus sequence generation, by iVar 1.3.1^24^. Only positions covered by over 85% of samples with a depth of at least 20 reads are included in the analysis, totaling 25,611 positions. To avoid confounding by the variability of sequencing depth and coverage, variant calling results were downsampled to a uniform depth of 20, in accordance with the method presented in Zhao et al.^4^. Annotations were performed using the Ensembl COVID-19 portal; global occurrence statistics are sourced from the RCoV19 database^25^.

### Genomic feature annotation and statistical analysis

#### Classification of mutational origin and rationale

The intra-host mutational landscape of SARS-CoV-2 is composed of two distinct classes: major, or clonal mutations, and minor mutations. Major mutations are “fixed”, with little to no other alleles in the host’s viral population, while the minor mutations often comprise a smaller fraction of the observed alleles^5^. For our data, we selected 90% allelic fraction as a reasonable cutoff between mutational classes (Figures 2). Longitudinal studies found that intra-host variability is mostly confined to minor, uncommon, nonsynonymous mutations; however, this criterion is not sufficiently accurate as a classifier^4^. To obtain a set of mutations that is highly enriched for iSNVs, we selected rare mutations that did not appear in any adjacent viral sequence on the phylogenetic tree. This is a strict criterion, but it is also the highest in specificity and aligns with the findings of prior longitudinal studies^4^ (Figure 4). Major mutations (present in over 90% of sequencing reads) represent the dominant viral population and are therefore mostly advantageous and persistent across hosts, while minor mutations are not persistent, with lower chances of transmission and more volatility (Figure 3).

**Figure 2:**
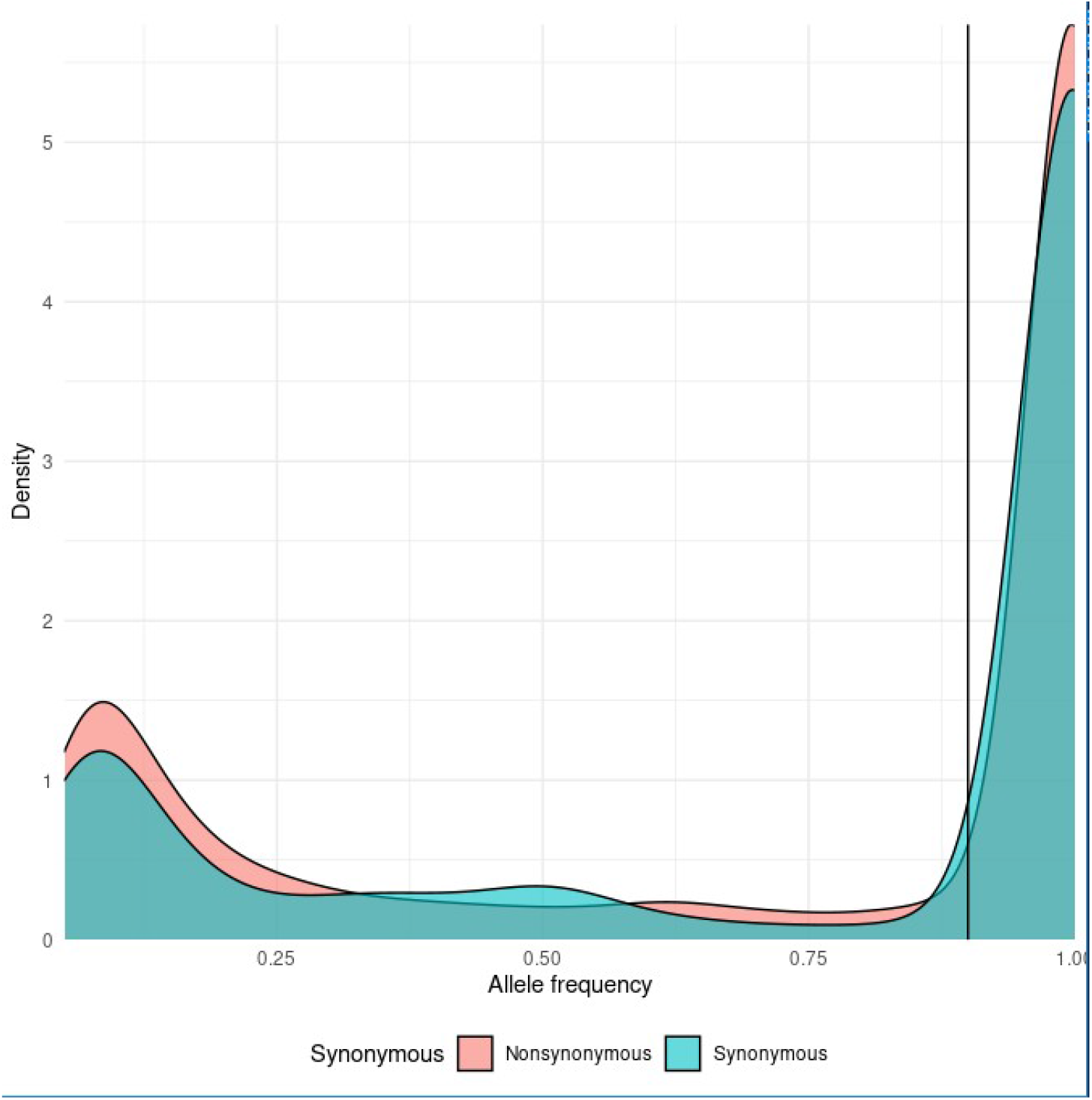
The distribution of allele frequencies for synonymous (cyan) and nonsynonymous (orange) mutations. The mutation class threshold (90%) is marked by a vertical line.

**Figure 3:**
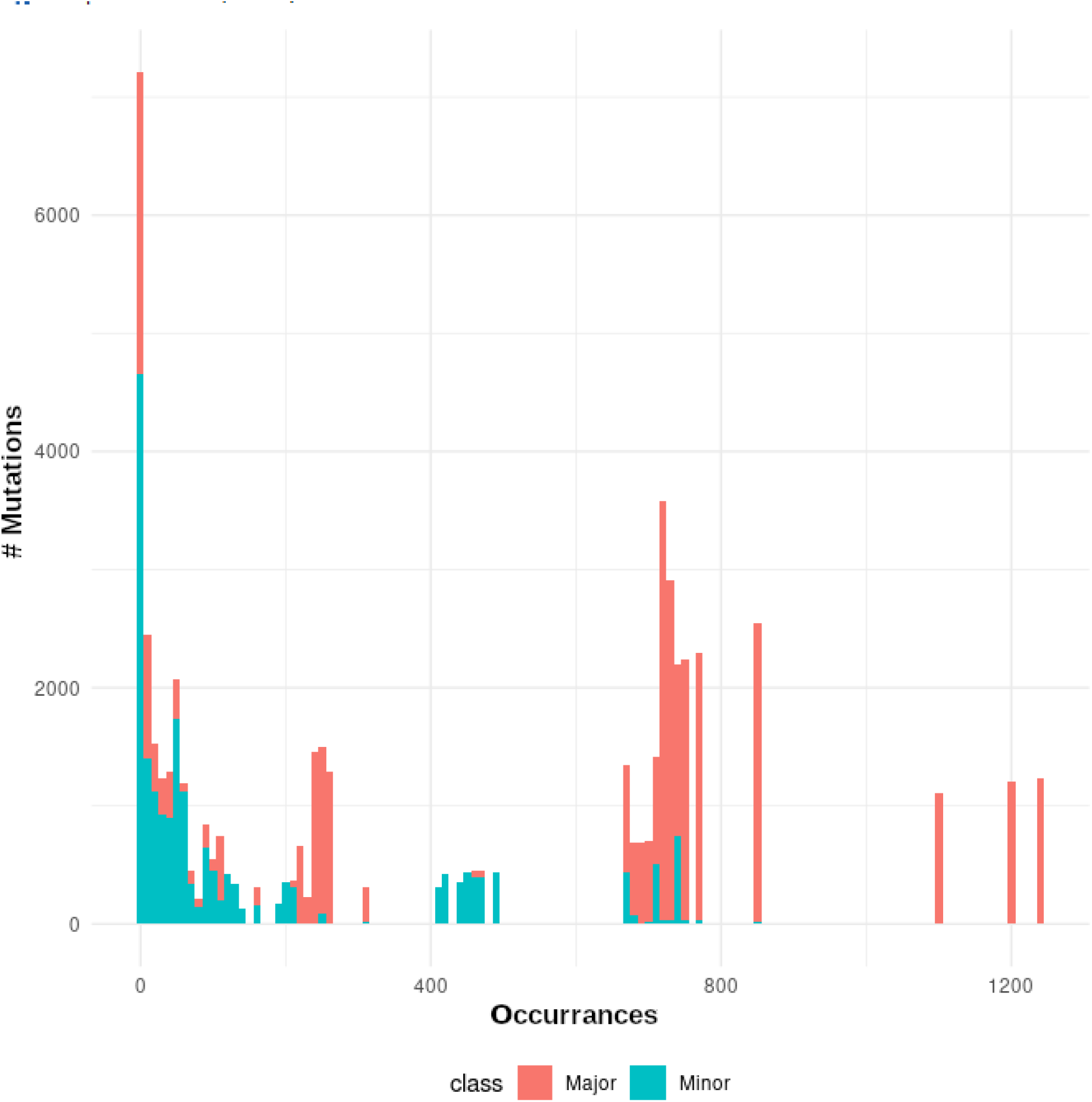
A histogram of mutations by the number of occurrences in the cohort, colored according to their assigned class.

**Figure 4:**
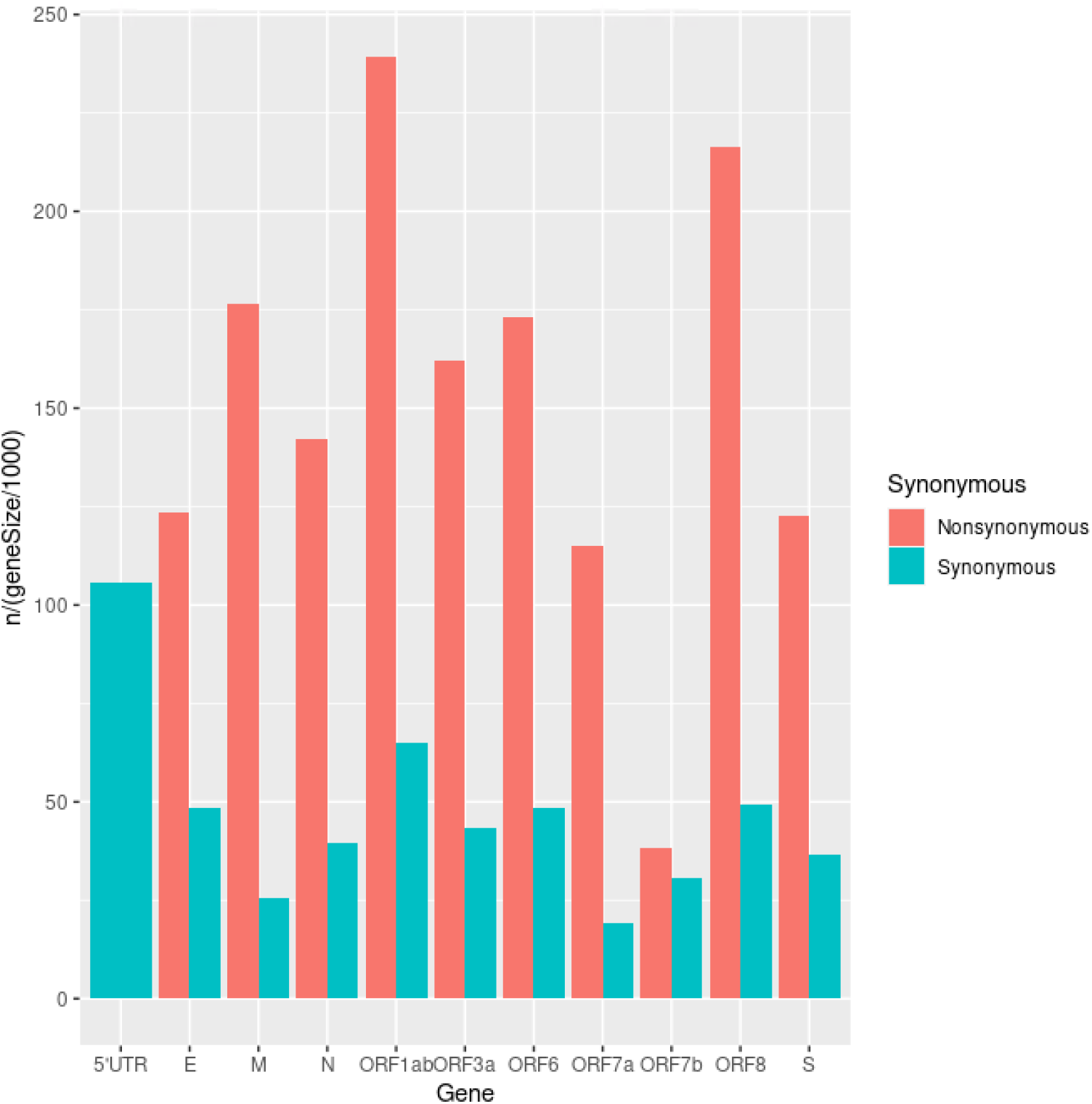
A bar plot of the number of iSNVs in each gene, corrected for gene length and colored according to type (non/synonymous).

**Figure 5:**
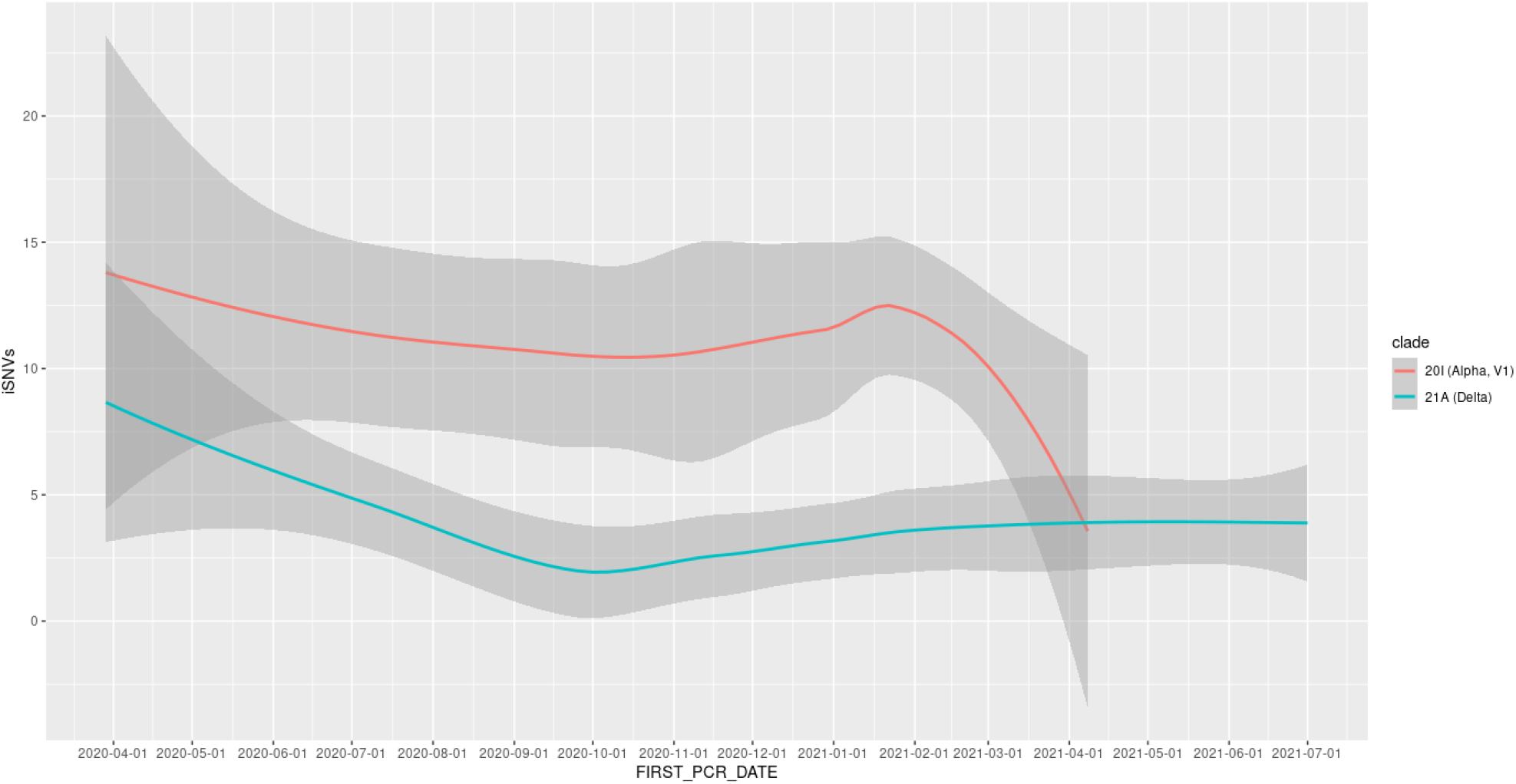
LOESS trend line depicting the change in intra-host diversity over time, by viral clade.

#### Statistical modeling

A generalized Poisson model was used for modeling intra-host variability and testing against individual clinical parameters. The amount of iSNVs, as determined according to our methodology, was set as the dependent variable. The independent variables were selected and tested systematically from the available electronic health records data. To ensure that our results were not skewed by host-independent factors, we considered the date of collection and viral clade as possible confounders. Sequencing coverage remained negatively correlated to the number of overall iSNVs and was thus added to the model as an additional confounder.

## Data Availability

The electronic health records used in this study are confidential and protected by law. Other data used in this study is currently private.

## Limitations

The primary limitation of our study is the lack of multiple (longitudinal) viral samples from individual participants over the course of infection. Our method for classifying iSNVs relies on statistics from large-scale surveillance databases and the tendency of iSNVs to produce a small set of mutations, highly enriched in iSNVs. Furthermore, our samples were generally taken shortly after infection, while samples taken during late infection are expected to contain more iSNVs^4^, which may be more influenced by host-pathogen interactions.

### Ethics declaration

This study was approved by the MHS Institutional Review Board. Due to its retrospective design, informed consent was waived, and all identifying details of the participants were removed before computational analysis.

